# Efficacy of Balloon Pulmonary Angioplasty in Chronic Thromboembolic Pulmonary Disease Patients with Exercise Pulmonary Hypertension

**DOI:** 10.1101/2024.02.19.24303059

**Authors:** Yutaro Naka, Takumi Inami, Kaori Takeuchi, Hanako Kikuchi, Ayumi Goda, Masaharu Kataoka, Takashi Kohno, Kyoko Soejima, Toru Satoh

## Abstract

**Background:** The efficacy of balloon pulmonary angioplasty (BPA) for chronic thromboembolic pulmonary disease (CTEPD) with or mild pulmonary hypertension (PH) or without PH remains unknown. Exercise pulmonary hypertension (Ex-PH) is associated with impaired exercise capacity and ventilatory efficiency, even under normalized pulmonary hemodynamics at rest. We hypothesized that patients with Ex-PH and/or hypoxemia would be candidates for BPA. We aimed to verify the prevalence and clinical profiles of Ex-PH and the effect of BPA on oxygenation and Ex-PH in patients with CTEPD with mean pulmonary arterial pressure (mPAP) < 25 mmHg.

**Methods:** We retrospectively reviewed 29 patients with CTEPD and mPAP < 25 mmHg at rest, who had undergone a cardiopulmonary exercise test with right heart catheterization (median age, 65 years; 38% male). Patients were divided into two groups: Ex-PH, defined as a cardiac output slope (mPAP/CO slope) > 3.0, and non-Ex-PH.

**Results:** Overall, six patients had mild PH (mPAP: 21–24 mmHg), and 16 and 13 were assigned to the Ex-PH and Non-Ex-PH groups, respectively. There were no significant differences in the clinical parameters, including hemodynamics at rest, blood gas analysis, and 6-minute walk distance, between the Ex-PH and Non-Ex-PH groups. Among the 16 patients with Ex-PH and/or long-term oxygen therapy (LTOT), BPA improved the World Health Organization-functional class (WHO-FC) and PaO_2_ in association with a decrease in the mPAP/CO slope. All nine patients discontinued LTOT after BPA. No significant complications were observed during each BPA session.

**Conclusions:** Ex-PH was common among patients with CTEPD and mPAP < 25 mmHg. BPA can improve symptoms, oxygenation, and exercise hemodynamics in patients with CTEPD and Ex-PH and/or hypoxemia.

**What is Known?:**

- BPA has been recommended for patients with non-operable CTEPH.
- Although there is still a small body of evidence, BPA for patients with CTEPD with mild PH (mPAP < 25 mmHg) or without PH can safely improve symptoms.
- The prevalence of Ex-PH in CTEPD patients with or without PH is unknown.

**What the Study Adds?:**

- Approximately 50% of CTEPD patients with mild PH or without PH had Ex-PH.

In patients with CTEPD with mPAP < 25 mmHg, BPA improves exercising hemodynamics, such as the mPAP/CO slope, which could be a parameter to determine the indication for BPA.

Graphical Abstract.
The distribution of exercise pulmonary hypertension (Ex-PH) in patients with chronic thromboembolic pulmonary disease (CTEPD) with mild pulmonary hypertension (PH) or without PH, and efficacy of balloon pulmonary angioplasty (BPA) for CTEPD with Ex-PH and/or hypoxemia. Blue person symbols mean Ex-PH, and white person symbols mean non-Ex-PH.

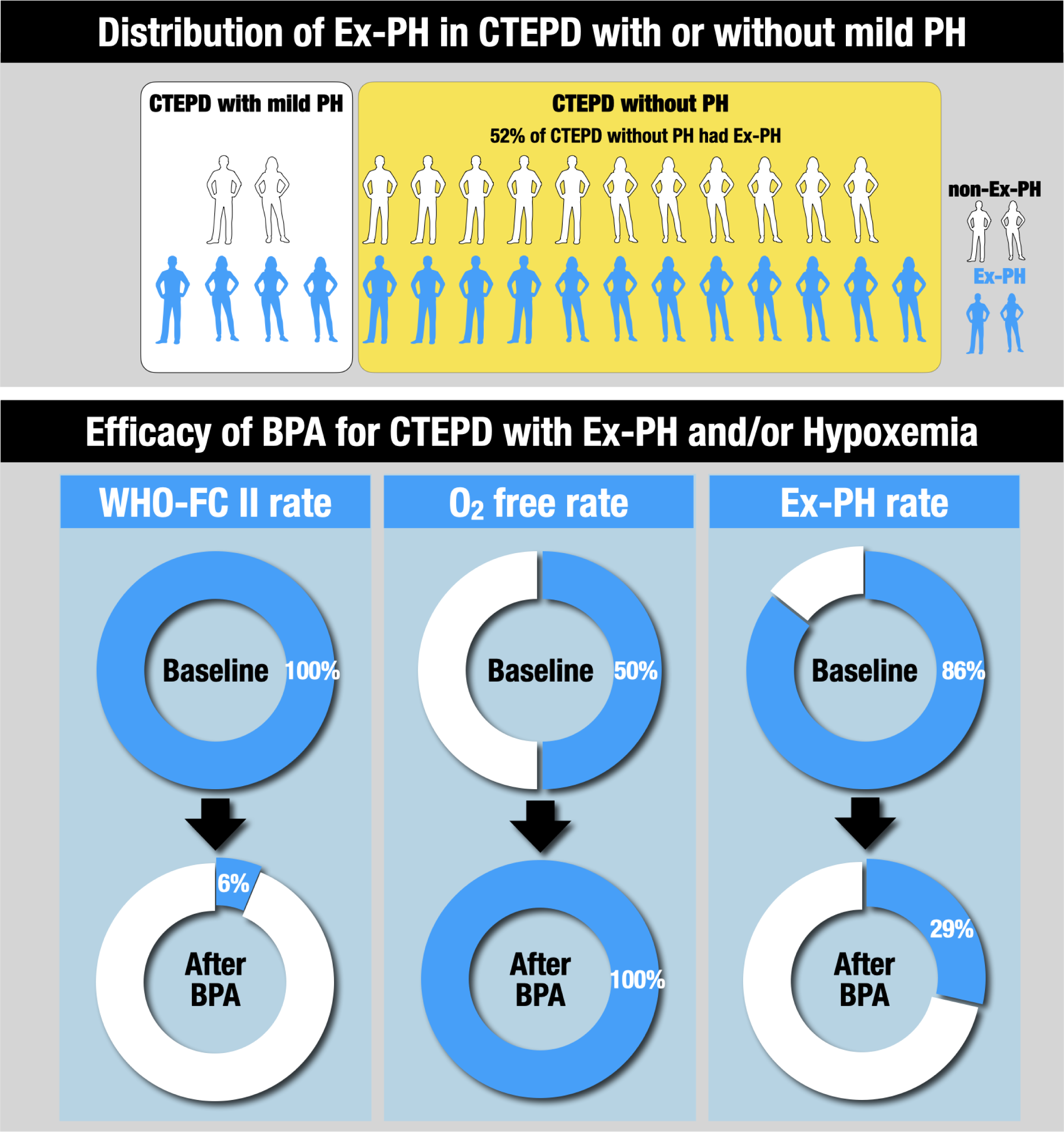

## Introduction

Recent clinical guidelines on the diagnosis and treatment of pulmonary hypertension (PH) have introduced the term chronic thromboembolic pulmonary disease (CTEPD) with or without PH, acknowledging the presence of similar symptoms, perfusion defects, and organized fibrotic obstructions in patients with or without PH at rest.^1^ In addition, a new definition of PH was recommended by lowering the mean pulmonary artery pressure (mPAP) threshold from 25 to 20 mmHg,^1^ based on the findings of pulmonary hemodynamics in healthy participants^2^ and the prognostic impact of borderline mPAP in patients with systemic sclerosis.^3^ Currently, research on the characteristics, natural history, and therapeutic strategies of CTEPD patients with mild PH and those without PH is required.

Balloon pulmonary angioplasty (BPA) has been upgraded to a class I recommendation in the therapeutic algorithm of chronic thromboembolic pulmonary hypertension (CTEPH), a preferred term in CTEPD with PH. BPA improves prognosis, hemodynamic parameters, physical function, and oxygenation in patients with inoperable CTEPH.^4–6^ There is still debate over which patients with CTEPD patients with mild PH and without PH should be treated with BPA.^7–9^ Exercise pulmonary hypertension (Ex-PH), defined as an mPAP/cardiac output (CO) slope >3 mmHg/L/min between rest and exercise,^1^ has been reported to be associated with impaired exercise capacity and ventilatory efficiency as well as a higher incidence of cardiovascular events, even under normalized pulmonary hemodynamics at rest.^10–12^ However, few studies have evaluated the benefits and safety of BPA in patients with CTEPD with Ex-PH, especially in treatment-naïve cases.^7, 8^ We hypothesized that patients with Ex-PH could be candidates for BPA among those with mild or no PH.

This study aimed to verify (1) the prevalence and clinical profiles of Ex-PH in patients with CTEPD with mild PH or without PH, (2) the effect of BPA on the pulmonary vascular response during exercise in Ex-PH, and (3) the long-term clinical outcomes of conservative management in non-Ex-PH.

## Methods

### Study Design and Participants

This was a retrospective longitudinal observational study. Twenty-nine patients with CTEPD and mPAP <25 mmHg who underwent a cardiopulmonary exercise test with right heart catheterization (CPET-RHC) were consecutively enrolled at a single Japanese university hospital (Kyorin University Hospital) between March 2014 and September 2021. CTEPD was diagnosed by the presence of organized pulmonary thromboembolism on contrast-enhanced lung computed tomography, perfusion lung scintigraphy, and pulmonary angiography. Right heart catheterization (RHC) was performed to confirm mPAP <25 mmHg and pulmonary artery wedge pressure (PAWP) <15 mmHg at rest. We excluded patients who had been treated with BPA-and/or PH-targeted drugs before enrollment. After a comprehensive evaluation using blood tests, pulmonary function tests, and echocardiography, patients with collagen vascular disease, chronic respiratory disease, left heart abnormality, or other systemic diseases were excluded. This study complied with the Declaration of Helsinki and was approved by the Committee for Clinical Studies and Ethics of Kyorin University School of Medicine. Written informed consent was obtained from each patient before participation in the study.

### Clinical Variables

The patients’ clinical variables were obtained from their electronic medical records, and we collected data on the patients’ demographics (age, sex), previous history of venous thromboembolism (VTE), treatment (long-term oxygen therapy [LTOT], anticoagulation), functional capacity (World Health Organization-functional class [WHO-FC], six-minute walk distance [6-MWD]), laboratory and echocardiographic data, pulmonary arterial angiography, RHC at rest and during exercise, and cardiopulmonary exercise test (CPET). Based on the pulmonary angiography findings, the number of organized thrombotic lesions was calculated as the cumulative number of weighted proximal lesions (segmental branch lesion, 1 point; subsegmental branch lesion, 0.5).

### RHC and CPET-RHC

We performed RHC using a 6-French double-lumen flow-directed balloon-tipped Swan-Ganz catheter (Harmac Medical Products, Inc., Buffalo, NY, USA) via the transjugular approach as previously described.^11–13^ We determined the CO using the direct Fick method with assumed oxygen consumption. Based on the new definition of PH in European Society of Cardiology/European Respiratory Society (ESC/ERS) guidelines 2022^1^, patients who were previously referred to as having chronic thromboembolic disease (mPAP <25 mmHg) were divided into two categories 1) CTEPD with mild PH (20 < mPAP < 25 mmHg) and 2) CTEPD without PH (mPAP ≤20 mmHg).

CPET-RHC was performed using Cpex-1^®^ (Inter-Reha, Tokyo, Japan), as described previously.^11^ Briefly, using the ramp protocol, we performed an incremental symptom-limited exercise test with an electromagnetically braked cycle ergometer (Nuclear Imaging Table with Angio Ergometer; Lode, Groningen, the Netherlands). Hemodynamics were measured during exertion by using a catheter with a tip in the trunk of the pulmonary artery. The mPAP-CO during exercise was plotted using multipoint plots. We defined Ex-PH as an mPAP/CO slope >3.0, according to the ESC/ERS guidelines 2022,^1^ and the enrolled patients were categorized into Ex-PH and non-Ex-PH groups (Figure 1).

**Figure 1.**
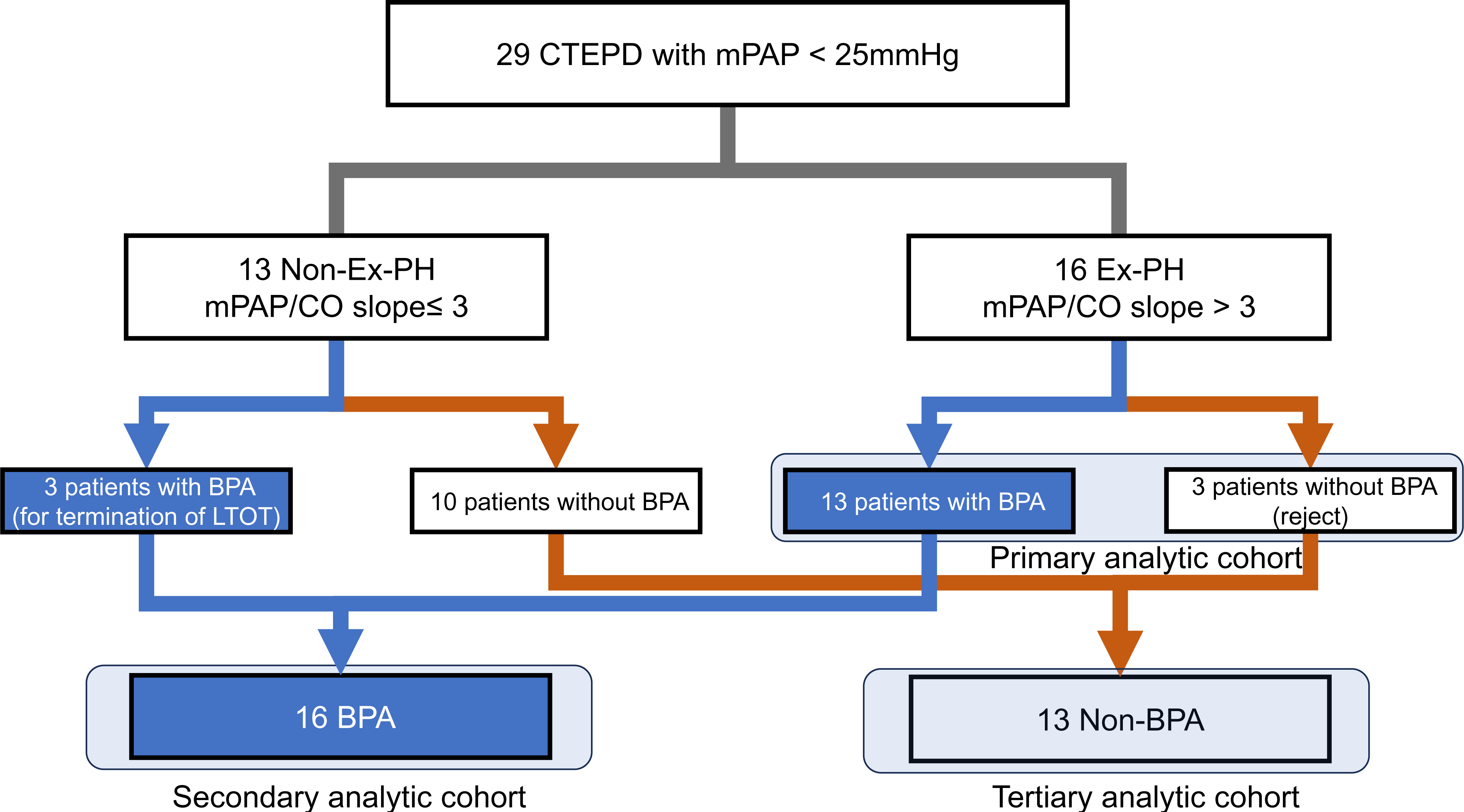
The study cohort flowchart. BPA, balloon pulmonary angioplasty; CTEPD, chronic thromboembolic pulmonary disease; CPET-RHC, cardiopulmonary exercise test with right heart catheterization; Ex-PH, exercise pulmonary hypertension; LTOT, long-term oxygen therapy; mPAP/CO slope, mean pulmonary arterial pressure/ cardiac output slope; PH, pulmonary hypertension.

### Indication and Procedure of BPA

Based on whether the enrolled patients were treated with BPA, they were categorized into BPA and non-BPA groups (Figure 1). BPA was indicated when the patient had exertional dyspnea due to Ex-PH or hypoxemia caused by organized thrombus occlusion, as follows (all items fulfilled): 1) mPAP/CO slope >3.0 or receiving LTOT; 2) WHO-FC >II; and 3) understanding of the procedures and possible complications of BPA and submission of informed consent. Although we considered the indications for pulmonary endarterectomy (PEA) in all patients, they underwent BPA for the following reasons: 1) peripheral lesion type, 2) inoperability due to the high risk of general anesthesia or poor physical condition, and 3) patient refusal to undergo PEA. Details of the interventional procedures have been described^14^. Briefly, all procedures were performed via the femoral approach under local anesthesia. A 7-French guide catheter was inserted through the sheath and a 0.014-inch guidewire was passed through the target lesion. Approximately 1.2–8.0 mm monorail balloons were used to dilate the target lesions. Pulmonary arterial angiography of the lungs, including the target lesions, was performed before each session to select and determine the target lesions. A 0.014-inch guide wire was used with a pressure catheter (Navvus II^®^: ACIST, Minnesota, USA) by passing across the target lesion, as described previously.^15^ The pulmonary artery pressure (PAP) proximal and distal to the target lesion and the ratio of the two pressures were measured using a pressure catheter. The balloon dilatation endpoint, in terms of effectiveness, was to finish the dilatation when the pressure gradient ratio of the distal to proximal pressures across the target lesion detected by the pressure catheter was >0.7. All lesions that were deemed catheterizable were treated whenever possible.

PH-targeted medications were not administered to patients in this cohort during the observation period. Data on BPA-related complications, such as pulmonary injury, contrast-induced nephropathy, the need for hemodialysis, extracorporeal membrane oxygenation or mechanical ventilation, and death, were collected from patients who underwent BPA. RHC was performed 6–12 months after BPA. Termination of the LTOT criteria after BPA was defined as PaO_2_ >60 mmHg at rest and peak oxygen consumption (VO_2_). Within 1 d of the corresponding RHC, laboratory data and 6-MWD were assessed. In patients without BPA, echocardiography was serially performed for 2 years after enrollment.

### Statistical Analysis

Data are presented as medians (interquartile ranges), and categorical variables are expressed as numbers and percentages. The Shapiro–Wilk test was used to assess the normality of the data distribution. Significant differences were determined using the Mann–Whitney U test and Wilcoxon matched-pair signed-rank test, as appropriate. Differences in frequencies were analyzed using Fisher’s exact test or the chi-squared test, as appropriate. Hemodynamic parameters during exercise were compared between the Ex-PH and non-Ex-PH groups using a two-way repeated-measures analysis of variance (ANOVA) and Sidak’s multiple comparison test, as appropriate. The analysis workflow is as follows (Figure 1): 1) as primary analysis, clinical characteristics, exercise capacity, laboratory, echocardiographic, hemodynamics (at rest and during exercise), and CPET parameters were compared between the Ex-PH and non-Ex-PH groups; 2) as secondary analysis, to evaluate the impact of BPA in the CTEPD patients, functional capacity, termination of LTOT, oxygenation, and hemodynamics (at rest and during exercise) were compared between baseline and after BPA, and clinical events (all-cause mortality and heart failure hospitalization) were evaluated; 3) as tertiary analysis, to verify the long-term clinical outcomes in the CTEPD patients without BPA treatment, serial echocardiography measurements and clinical events were evaluated. Statistical significance was set at p <0.05. All statistical analyses were performed using GraphPad Prism version 10.1.0 (GraphPad Software, Boston, MA, USA).

## Results

### Comparison of Parameters between non-Ex-PH and Ex-PH

Among the enrolled 29 patients (62.1% of female, median age of 64 [55–69]), 6 (20.7%) were categorized as CTEPD with mild PH (mPAP: 21–24 mmHg), and 2 (6.9%) were categorized as precapillary PH based on the definition of the ESC/ERS 2022 guidelines (pulmonary vascular resistance [PVR] >2 Wood/U and mPAP >20 mmHg)^1^ (Graphical Abstract and Table 1).

**Table 1.**
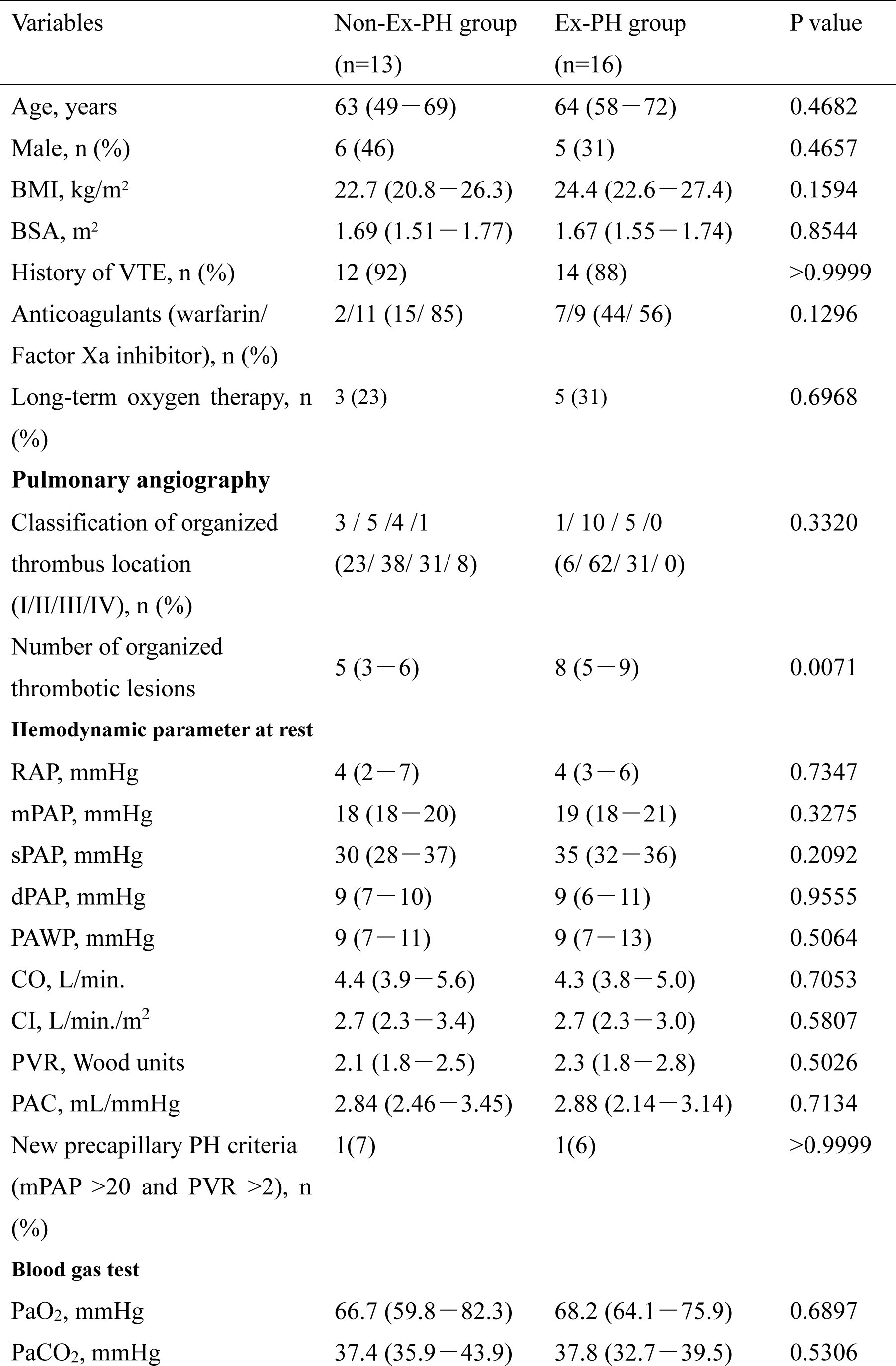
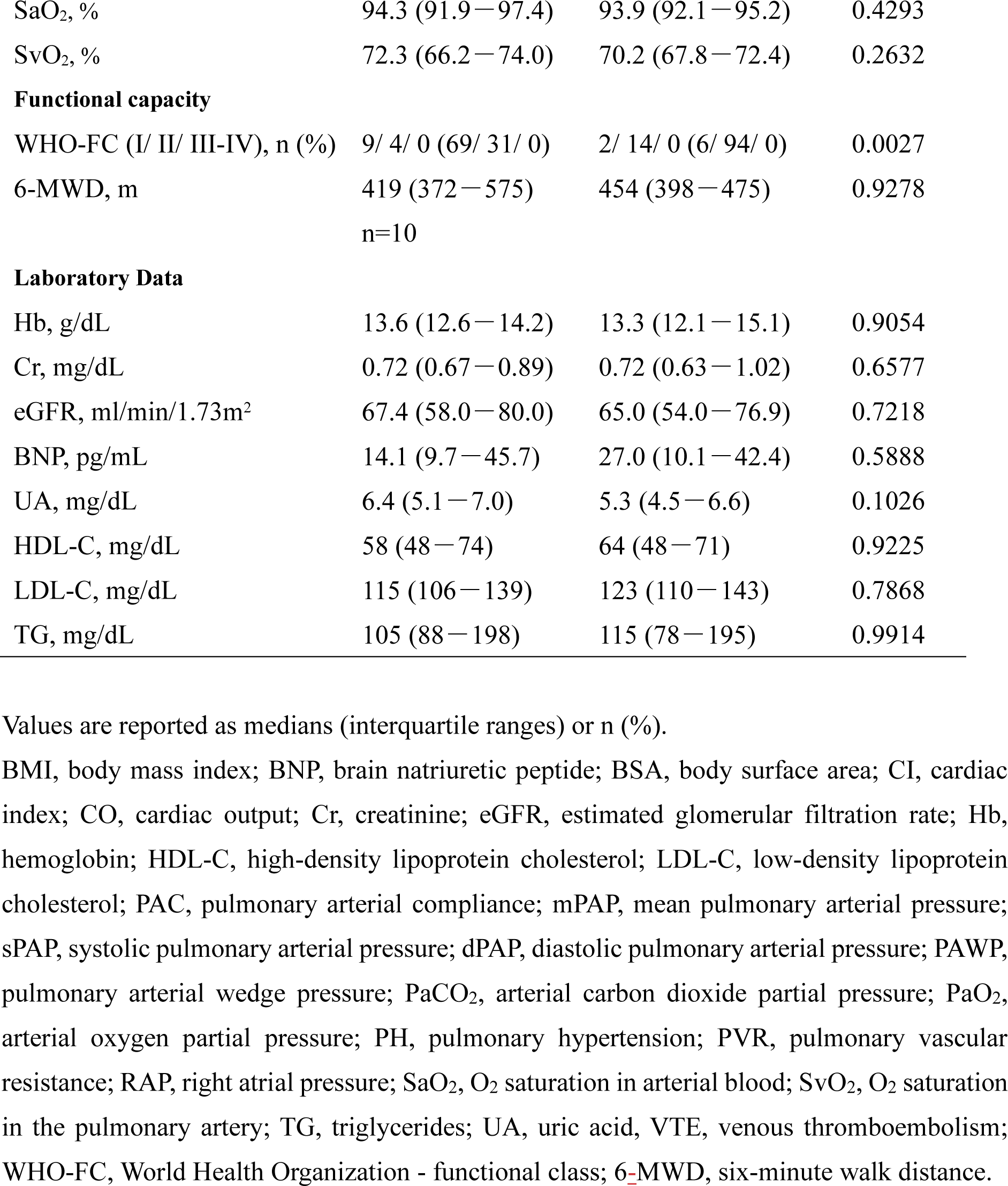
Baseline characteristics.

Overall, 13 (44.8%) and 16 (55.2%) patients were classified into non-Ex-PH and Ex-PH groups, respectively. Among 23 without PH, 12 (52.2%) were categorized as Ex-PH, respectively. Patient characteristics are detailed in Tables 1 and 2. No significant differences were observed in the patient demographics (age, sex, and history of VTE), LTOT use, classification of organized thrombus location, hemodynamic parameters at rest, 6-MWD, and laboratory and echocardiographic data between the non-Ex and Ex-PH groups. In the non-Ex-PH group, the number of organic thrombotic lesions was lower (5 [3–6] vs. 8[5–9], p <0.0071) and WHO-FC was lower. Among the CPET-RHC parameters (Figure 2), the mPAP-CO slope (2.4 [2.1–2.6] vs. 4.6 [3.6–6.2] mmHg, p <0.0001) and mPAP at peak exercise (35 [30–42] vs. 47 [40–51] mmHg, p=0.0053) were lower in the non-Ex-PH group than in the Ex-PH group. There was no significant difference in peak VO_2_ (15.4 [11.1–18.8] vs. 14.4 [12.8–15.5] mL/min/kg, p = 0.5890) and minute ventilation (VE) / carbon dioxide output (VCO_2_) slope (35.7 [32.8–49.2] vs. 36.6 [32.3–44.4], p = 0.7407) between the non-Ex-PH and Ex-PH groups (Supplemental Table).

**Figure 2.**
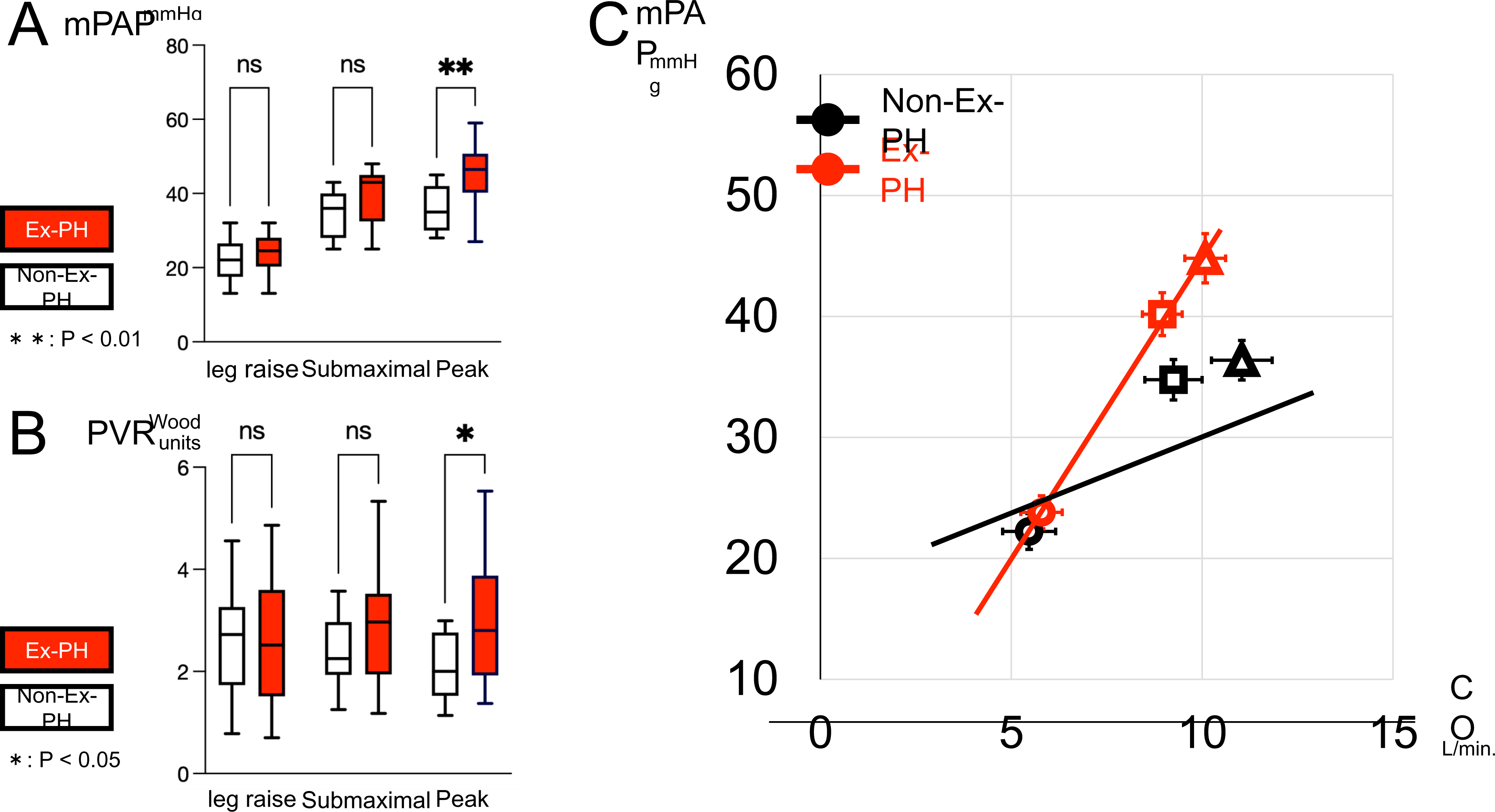
Comparison of exercising hemodynamic changes in the mean pulmonary arterial pressure (mPAP) **(A)**, the pulmonary vascular resistance (PVR) **(B)**, and mPAP/ cardiac output (CO) slope **(C)** between non-exercise pulmonary hypertension (Non-Ex-PH) group and Ex-PH group. In Figure 2A and B, bars denote medians, boxes denote interquartile ranges, and whiskers denote ranges excluding statistical outliers (circles) (>1.5 box lengths from either the 25^th^ or 75^th^ percentiles). In Figure 2C, values represent mean (symbol) ± SEM (bar). The circle symbol means the value at leg raise. The square symbol means the value at the submaximal. The triangle symbol means the value at peak oxygen consumption. The lines mean mPAP/ CO slope.

**Table 2.**
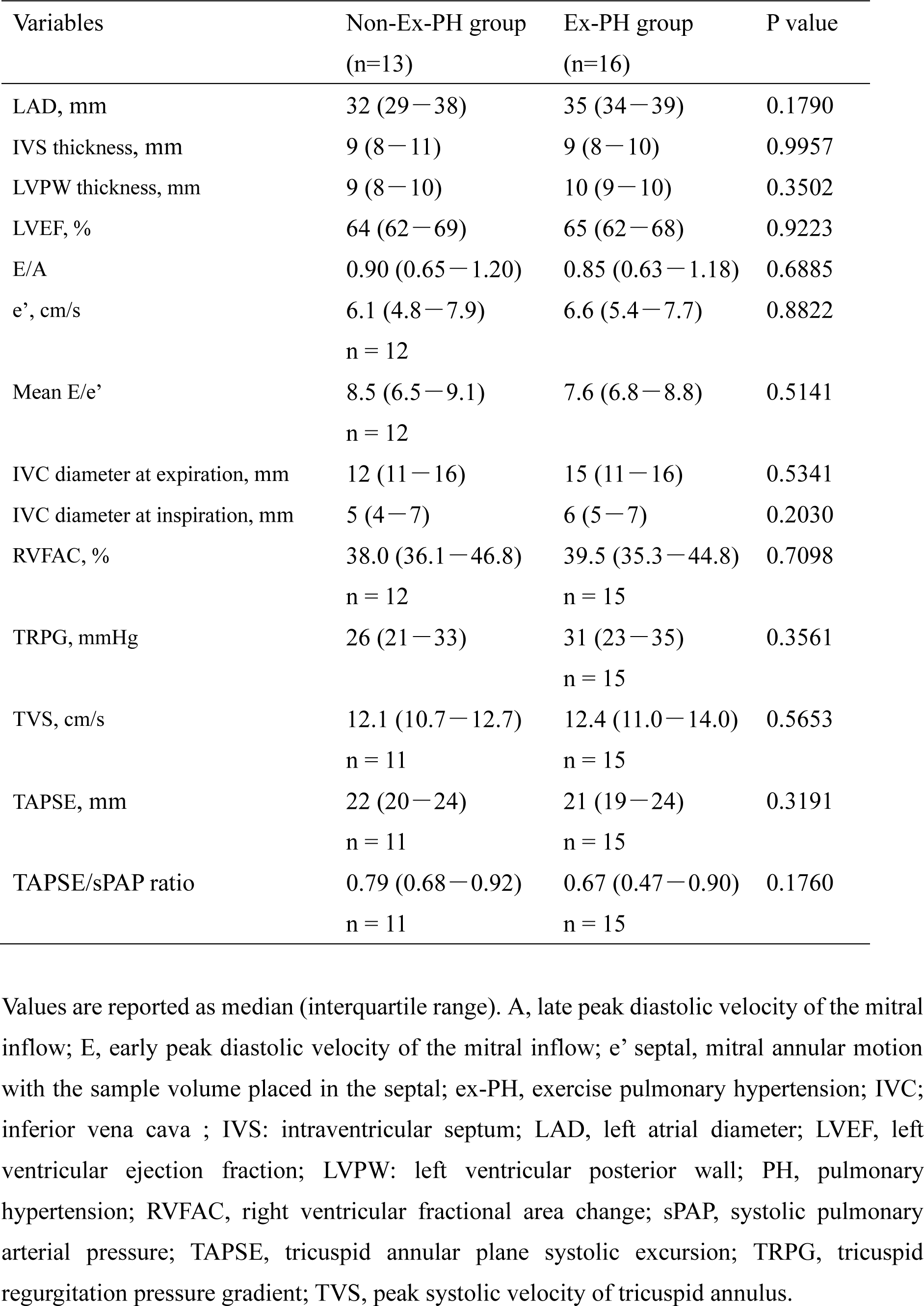
Echocardiographic findings.

### Hemodynamics, Functional Capacity, and LTOT use after BPA

Among the 16 patients with Ex-PH, 13 consented to undergo BPA and 3 did not (Figure 1). All three non-Ex-PH patients treated with LTOT underwent BPA to terminate LTOT. Thus, BPA was performed on 16 patients with CTEPD in this cohort. The number of dilated vessels per patient was 12.0 (8.0–13.5), with 2 (2–3) sessions performed per patient. There were no significant complications, such as pulmonary injury, contrast-induced nephropathy, or the need for mechanical circulation support or ventilation, during each BPA session.

Hemodynamic parameters at rest, oxygenation, and exercise capacity before and after BPA are shown in Table 3. Resting mPAP and systolic PAP improved after BPA. The PVR, CO, and pulmonary arterial compliance (PAC) did not change after BPA treatment. WHO-FC (I: II = 0:17 to 16:1, p <0.0001), and oxygenation at rest (PaO_2_:66.6 [62.0–76.5] to 77.4 [67.9–90.4], p = 0.0181) improved after BPA, and all eight patients discontinued LTOT (Figure 3).

**Figure 3.**
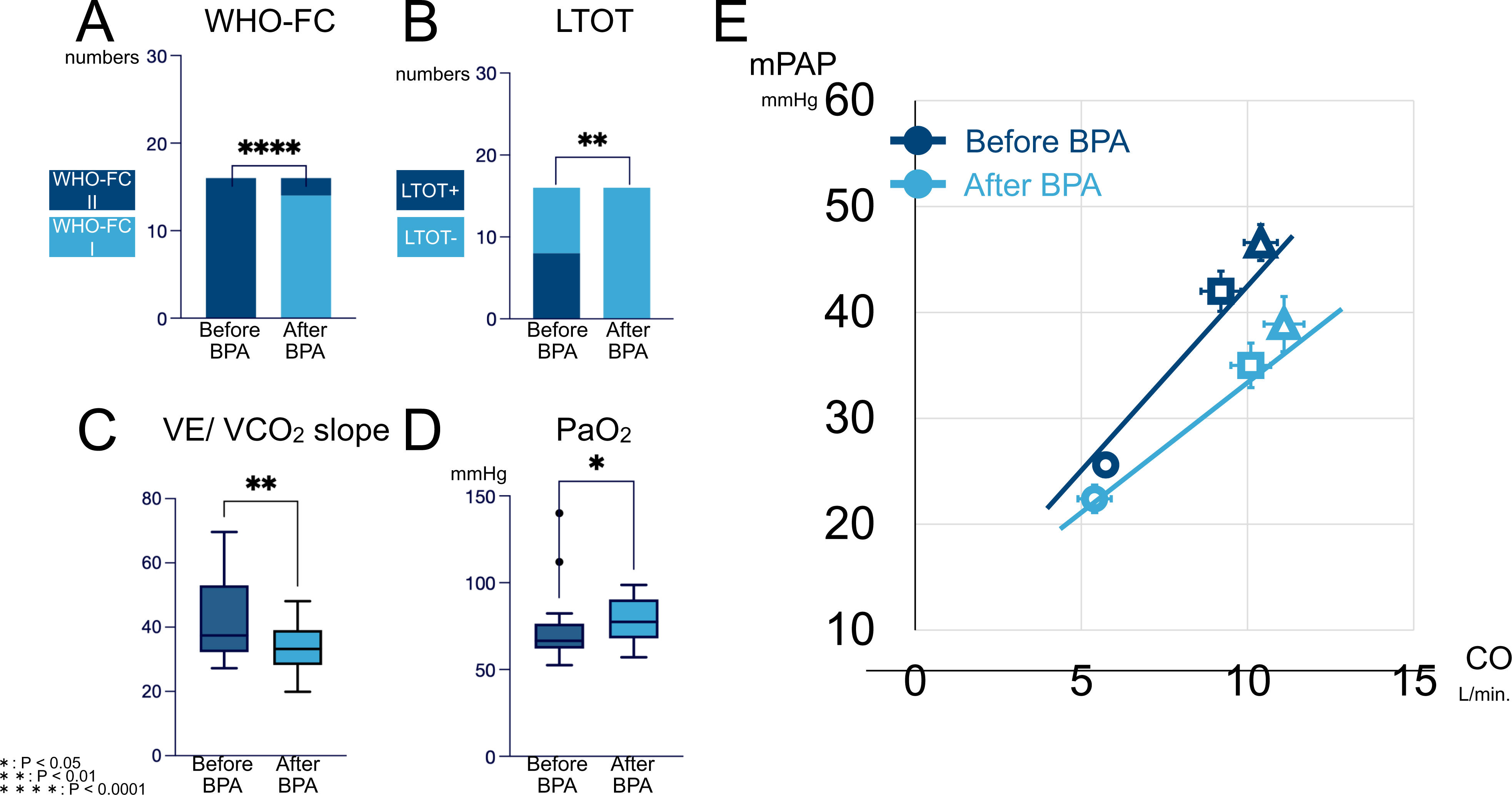
Efficacy of balloon pulmonary angioplasty (BPA) for CTEPD with exercise pulmonary hypertension (Ex-PH) and/or hypoxemia. World Health Organization-functional class (WHO-FC; **A**) and use of long-term oxygen therapy (LTOT; **B**) before and after BPA. Values represent numbers in Figure 3A and B. Change in arterial oxygen partial pressure (PaO_2_; **C**) and minute ventilation (VE) / carbon dioxide output (VCO_2_) slope (**D**) before and after BPA. In Figures 3C and D, bars denote medians, boxes denote interquartile ranges, and whiskers denote ranges excluding statistical outliers (circles) (>1.5 box lengths from either the 25^th^ or 75^th^ percentiles). mPAP/CO slope (**E**) before and after BPA. In Figure 3E, values represent mean (symbol) ± SEM (bar). The circle symbol means the value at leg raise. The square symbol means the value at the submaximal. The triangle symbol means the value at peak oxygen consumption. The lines mean mPAP/CO slope.

**Table 3.** Efficacy of BPA for hemodynamic parameters at rest, oxygenation, and exercise capacity (n=16)

| Variables | Baseline | After BPA | P value |
| --- | --- | --- | --- |
| HR, bpm | 64 (58–68) | 68 (54–72) | 0.3557 |
| RAP, mmHg | 5 (4–7) | 4 (3–5) | 0.0835 |
| mPAP, mmHg | 20 (19–21) | 17 (14–19) | 0.0023 |
| sPAP, mmHg | 35 (33–36) | 30 (25–32) | 0.0020 |
| dPAP, mmHg | 9 (7–11) | 7 (6–10) | 0.1268 |
| PAWP, mmHg | 10 (8–12) | 8 (6–10) | 0.0851 |
| CO, L/min | 4.3 (3.9–5.1) | 4.2 (3.8–5.0) | 0.5533 |
| CI, L/min/m<sup>2</sup> | 2.7 (2.2–3.0) | 2.5 (2.3–3.1) | 0.5365 |
| PVR, Wood units | 2.2 (1.7–2.8) | 2.1 (1.4–2.7) | 0.2162 |
| PAC, mL/mmHg | 2.97 (2.33–3.40) | 3.12 (2.68–3.79) | 0.2114 |
| 6-MWD, m | 458 (380–520) | 488 (373–579) | 0.2769 |
| LTOT, n (%) | 8 (50) | 0 (0) | 0.0024 |

Values are reported as median (interquartile range). CI, cardiac index; CO, cardiac output; HR, heart rate; LTOT, long-term oxygen therapy; PAC, pulmonary arterial compliance; mPAP, mean pulmonary arterial pressure; sPAP, systolic pulmonary arterial pressure; dPAP, diastolic pulmonary arterial pressure; PAWP, pulmonary arterial wedge pressure; PVR, pulmonary vascular resistance; RAP, right atrial pressure; SpO<sub>2</sub>, saturation of percutaneous oxygen; 6-MWD, 6-minute walk distance.

The hemodynamic and cardiopulmonary functions of CPET-RHC before and after BPA are shown in Figure 3 and Table 4. Among the 16 patients who underwent BPA, two patients were unavailable for follow-up CPET-BPA because consent was not obtained. The peak work rate significantly increased (68 [50–90] to 70 [69–90] p = 0.0469), mPAP at peak significantly decreased after BPA (49 [42–51] to 40 [29–43] mmHg, p = 0.0005), CO at peak did not differ (10.5 [9.5–11.3] to 11.1 [9.3–12.7] L/min, p = 0.2676), and the mPAP/CO slope and VE/ VCO_2_ slope were reduced after BPA (mPAP/ CO slope: 4.1 [3.5–6.3] to 2.4 [1.3–4.1], p = 0.0067; VE/ VCO_2_ slope: 37.4 [32.2–53.1] to 33.2 (28.2–39.1), p = 0.0052). Four of 14 patients (29 %) had Ex-PH even after BPA (Graphical Abstract).

**Table 4.**
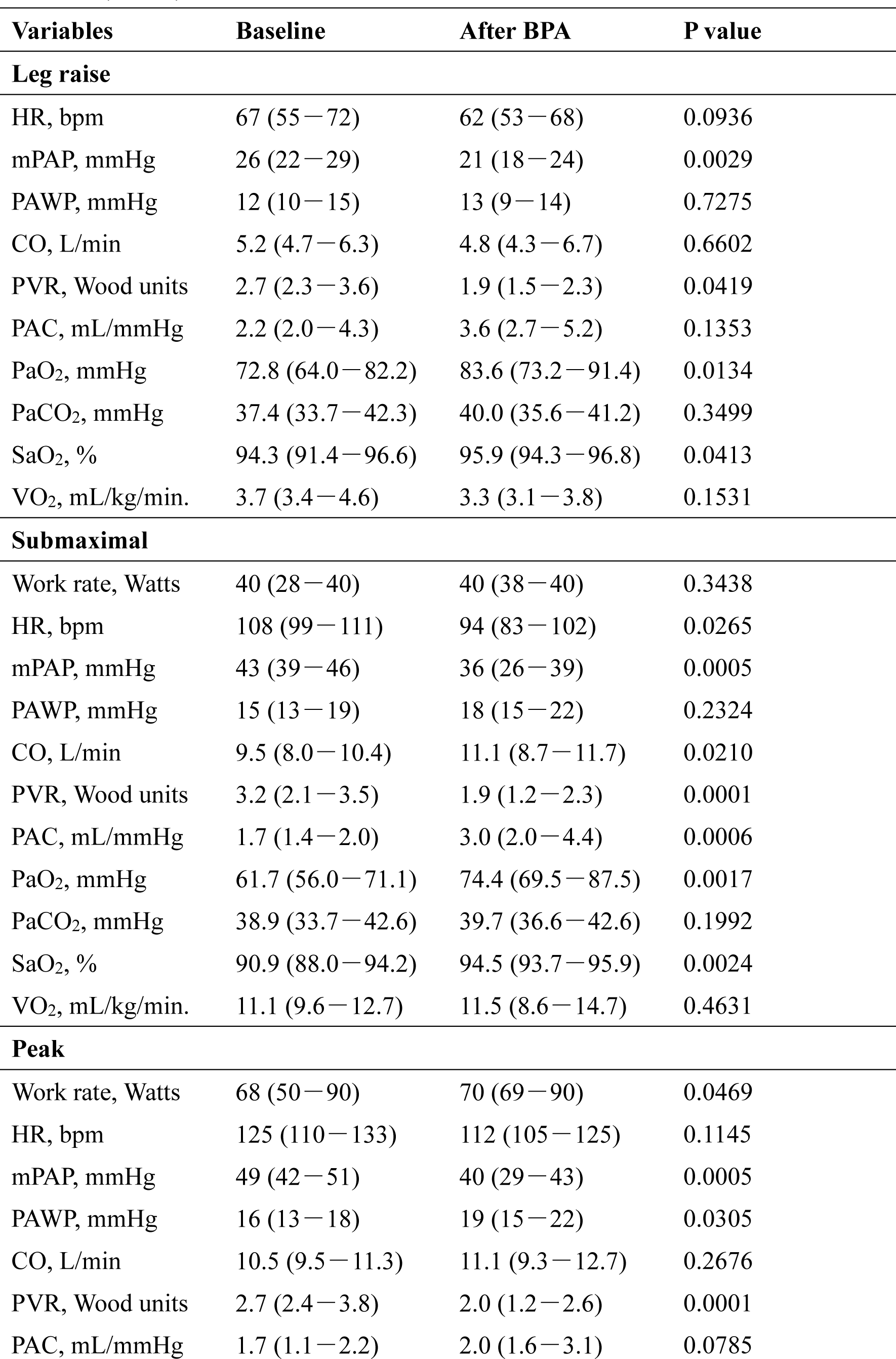
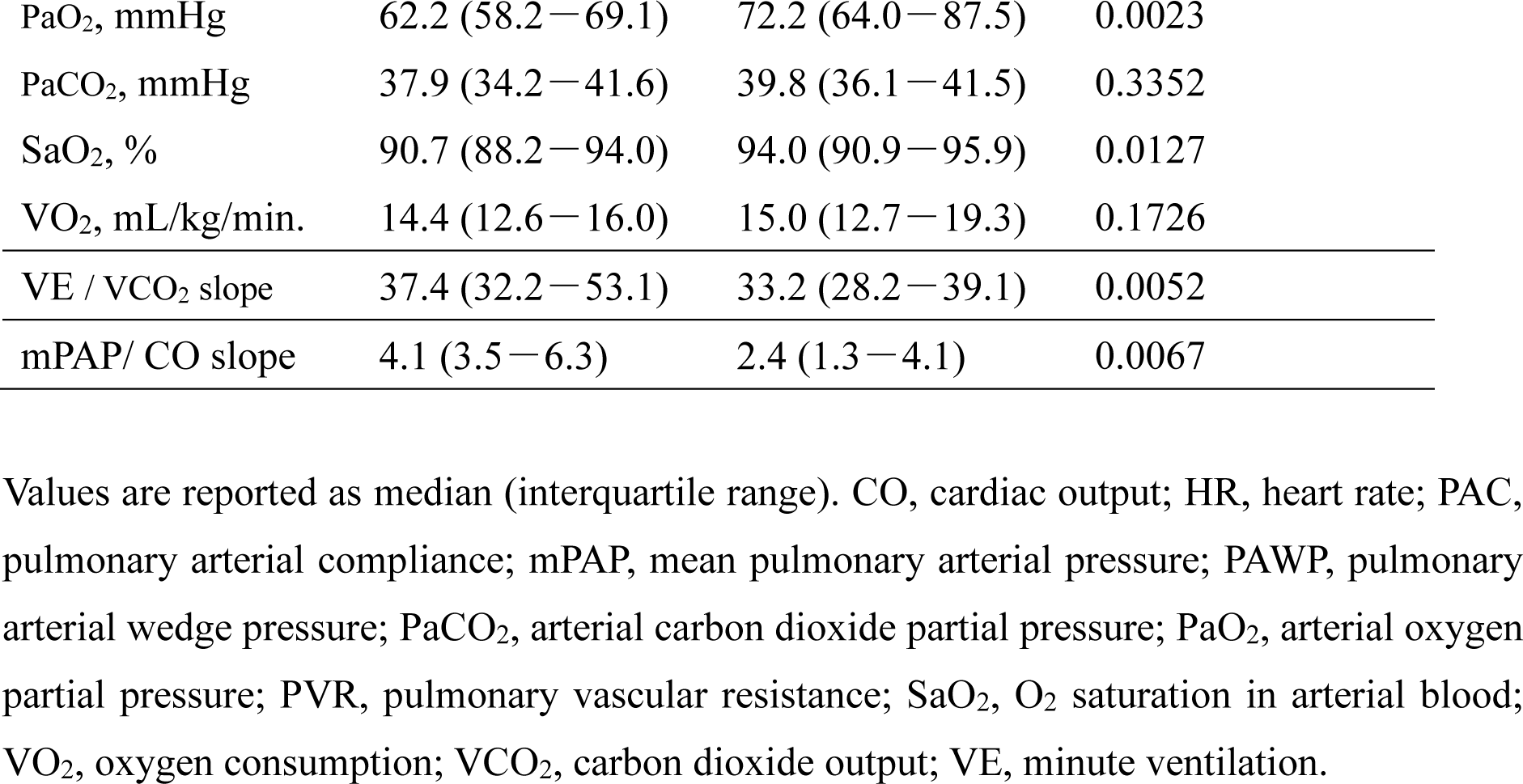
Efficacy of BPA for hemodynamic parameters and cardiopulmonary functions at CPET (n = 14)

Among the patients treated with BPA, there were no clinical events such as all-cause mortality, heart failure hospitalization, and recurrent VTE after BPA during the observational period (12.2 [7.4–12.9] months).

### Clinical events and echocardiography among patients without BPA

Among 12 of 13 patients without BPA (one patient was lost to follow-up), there were no clinical events such as all-cause mortality, heart failure hospitalization, or recurrent VTE during the observation period (3.3 [1.3–5.1]) years. None of the patients were treated with pharmacological interventions. There was no significant change in the tricuspid regurgitation pressure gradient (26 [21–32] to 25 [20–36], p = 0.478) during the interval between echocardiographic evaluations (3.2 [2.2–4.7] years).

## Discussion

This study revealed that Ex-PH was common among CTEPD patients with mild PH and without PH. Ex-PH could not be differentiated based on clinical findings other than CPET-RHC parameters, number of organized thrombotic lesions, and WHO-FC. Furthermore, based on the indication criteria for Ex-PH and/or deoxygenation, BPA significantly and safely improved symptoms, resting and exercising hemodynamics (i.e., mPAP at rest and mPAP/CO slope), and oxygenation.

### Frequency of Ex-PH among CTEPD with mild PH or without PH

Despite the introduction of Ex-PH (mPAP/CO slope >3) in the 2022 ESC/ERS guidelines,^1^ its frequency in patients with mild or no PH remains unclear. We previously reported that approximately half of the patients with CTEPH who improved to mPAP <25 mmHg after BPA had Ex-PH, which was related to impaired exercise capacity.^11^ In this study, in treatment-naïve CTEPD patients, 67% of patients with mild PH, as well as 52% of those without PH had Ex-PH. These findings suggest that Ex-PH is common in both treatment-naïve and post-BPA patients with CTEPD with mild or no PH.

Owing to the redefinition of PH by the ESC/ERS guidelines (i.e., lowering the mPAP threshold from 25 to 20),^1^ the proportion of CTEPD patients with PH has increased. In a prospective cohort study concerning post-pulmonary thromboembolism, Held et al. reported that with the change to this new definition, the number of cases diagnosed as CTEPD with PH increased from 17 to 21, a relative increase of 23.5%.^16^ Here, we demonstrated that among 29 treatment-naïve CTEPD patients with mPAP <25 mmHg, 20.7% corresponded to mild PH (21– 24 mmHg of mPAP). Notably, 6.9% patients were categorized as precapillary PH (PVR >2 wood units and mPAP >20 mmHg) in our cohort; the increase in precapillary PH by <u>the</u> new ESC/ERS definition was modest.

### Effectiveness and safety of BPA for CTEPD with mild PH and without PH

Recently, the efficacy of BPA in CTEPD with PH has been reported in numerous clinical trials. Two randomized clinical trials (RCTs), MR BPA^5^ and RACE,^6^ showed that BPA improved hemodynamics to a greater extent than medical therapy in patients with inoperable CTEPH. However, in these trials, CTEPD patients with mild PH of 21–24 mmHg of mPAP were not included, and eligibility for Ex-PH was not mentioned. Although therapeutic strategies for CTEPD with mild PH and without PH have not been established, there are a few observational studies, including ours, regarding the efficacy and safety of BPA.^7, 8^ In a small observational study involving 10 patients (35 BPA interventions), BPA improved symptoms, resting hemodynamics, and exercise hemodynamics^8^. The strength of our study is that we evaluated pulmonary hemodynamics (at rest and during exercise) and CPET parameters before and after BPA in all 29 naïve CTEPD patients. Notably, BPA improved the hemodynamics during exercise (mPAP/CO slope, mPAP, and PVR at peak exercise) and respiratory function (pO_2_ at rest and VE / VCO_2_ slope) with the discontinuation of LTOT. These findings suggest that even in patients with CTEPD and mildly impaired pulmonary hemodynamics, improvement of Ex-PH and ventilation-perfusion mismatch by BPA could lead to the discontinuation of LTOT. We propose that the mPAP/CO slope, an indicator of the Ex-PH diagnostic criteria, should be considered an indication of BPA. Because exercise intolerance in patients with CTEPD includes not only Ex-PH but also hypoxemia at rest and during exercise, hypoxemia requiring LTOT could be an indication for BPA. In the future, RCT are needed to assess the efficacy of interventions targeting Ex-PH or hypoxemia to discontinue LTOT.

There were several differences in improved parameters after BPA in our cohort (reduced mPAP) and in previous report<u>s</u> (reduced PVR and increased PAWP and PAC)^8^, which may be explained by differences in body size (body mass index: 24 vs. 28 kg/m^2^) and race, with and without PH-targeted drug treatment (treatment-naïve: 100 vs. 80%), history of VTE (94 vs. 40%), and BPA strategy (with and without pressure-guided). In our cohort, the resting mPAP decreased from 20 to 17 mmHg; however, the change in absolute value was small. Therefore, the mPAP may not be an appropriate endpoint for assessing the efficacy of BPA. Because of the improvements in symptoms, hemodynamics, and oxygenation observed in our observational studies, the ideal endpoint parameters of BPA for these categories should be clarified in future RCTs.

In the RACE Study, 22 of 52 patients (42%) in the BPA group had adverse events; in the MR BPA Study, 14 of 32 patients (44%) had complications of blood sputum, hemoptysis, or alveolar bleeding.^5, 6^ In a previous observational study, one in 10 patients with chronic thromboembolic pulmonary disease had BPA-related complications^8^. Notably, none of the patients in our study experienced BPA-related complications. Because hemodynamic severity has been associated with the occurrence of complications in previous trials and observational studies,^14, 17^ it is reasonable to assume that the frequency could be lower in CTEPD with mPAP <25 mmHg than in those with relatively higher mPAP. Although there are few reports on the efficacy and safety of PEA for CTEPD with borderline PH,^12^ it is necessary to discuss whether BPA or PEA is more appropriate as a first-line treatment considering its safety and efficacy.

Despite the beneficial effect of riociguat in Ex-PH patients after BPA,^18^ there is little evidence regarding the usefulness of PH-targeted drugs for CTEPD without PH. Because of the nature of microarteriopathy, there could be a possible efficacy of PH-targeted drugs in Ex-PH in CTEPD patients without PH. Notably, in the MR BPA study, BPA significantly improved oxygenation, but riociguat did not,^5^ suggesting that PH-targeted drugs, unlike BPA, might have no direct effect on ventilation-perfusion mismatch.

### CPET-RHC is useful for PPEI

Recently, close follow-up and care of patients with post-pulmonary embolism with persistent symptoms, exercise intolerance, and right heart dysfunction have been highlighted. The FOCUS study showed that patients with post-pulmonary embolism impairment (PPEI) were at higher risk of developing CTEPH.^19^ Recent worldwide and Japanese nationwide prospective registries have reported a history of acute pulmonary embolism in CTEPH patients of more than 60% in Europe, the United States, and other, and about 40% in Japan.^20^ The very high prevalence of history of acute pulmonary embolism in our cohort (more than 80%) could be explained by our selection bias due to detailed follow-up of post-pulmonary embolism patients with persistent dyspnea on exertion, which is similar to that in the FOCUS study.^19^ A systematic follow-up algorithm for PPEI in association with CPET-RHC could be useful for efficient detection of CTEPH. The clinical significance of therapeutic intervention in patients without Ex-PH or hypoxemia remains unknown because of insufficient evidence of their natural history to authorize a suitable risk-benefit to establish a recommendation for BPA.^21^ There were no clinical worsening events for approximately 3 years among patients without Ex-PH who were not treated pharmacologically or interconventionally. Whether these patients have a reduced quality of life remains unknown, and further studies are needed to evaluate the effects of PPEI screening and treatment on patient-centered outcomes.

### Limitations of this study

The major limitations were the small number of patients and the single-center, retrospective, and observational design of this study. The number of cases was small, and even if there were no statistically significant differences, it was difficult to make a definitive determination because of the lack of statistical power. Due to the nature of observational studies, it is not possible to establish a causal relationship. RCTs are needed to further investigate the safety and efficacy of BPA in CTEPD patients with mildly impaired pulmonary hemodynamics, as discussed previously.

## Conclusion

Ex-PH is common in patients with CTEPD with mild and without PH. BPA can safely improve symptoms, resting and exercising hemodynamics, and oxygenation in patients with CTEPD with Ex-PH and/or hypoxemia. Furthermore, individualized data accumulation based on academic experience is important for developing a comprehensive treatment concept for patients with CTEPD.

## Data Availability

I state that all data referred to in the manuscript are available.

## Non-standard Abbreviations and Acronyms

6-MWD: six-minute walk distance
BPA: balloon pulmonary angioplasty
CO: cardiac output
CPET-RHC: cardiopulmonary exercise test with right heart catheterization
CTEPH: chronic thromboembolic pulmonary hypertension
CTEPD: chronic thromboembolic pulmonary disease
Ex-PH: exercise pulmonary hypertension
LTOT: long-term oxygen therapy
mPAP: mean pulmonary artery pressure
OTLS: organized thrombotic lesion score
PAWP: pulmonary artery wedge pressure
PEA: pulmonary endarterectomy
PH: pulmonary hypertension
PPEI: post-pulmonary embolism impairment
PVR: pulmonary vascular resistance
RHC: right heart catheterization
VO_2_: oxygen consumption
VTE: venous thromboembolism
WHO-FC: World Health Organization-functional class

---

**Supplemental Table.**
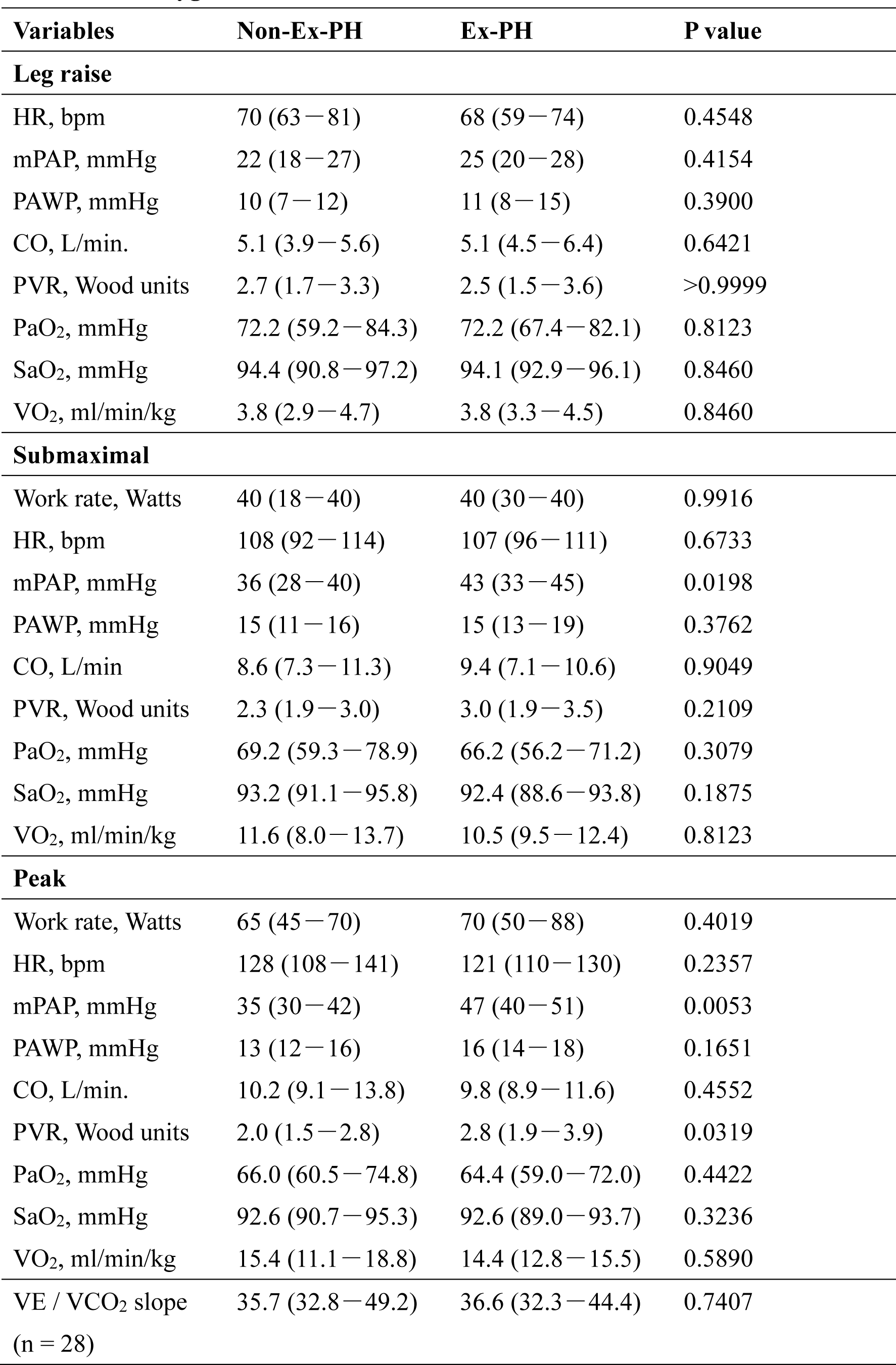
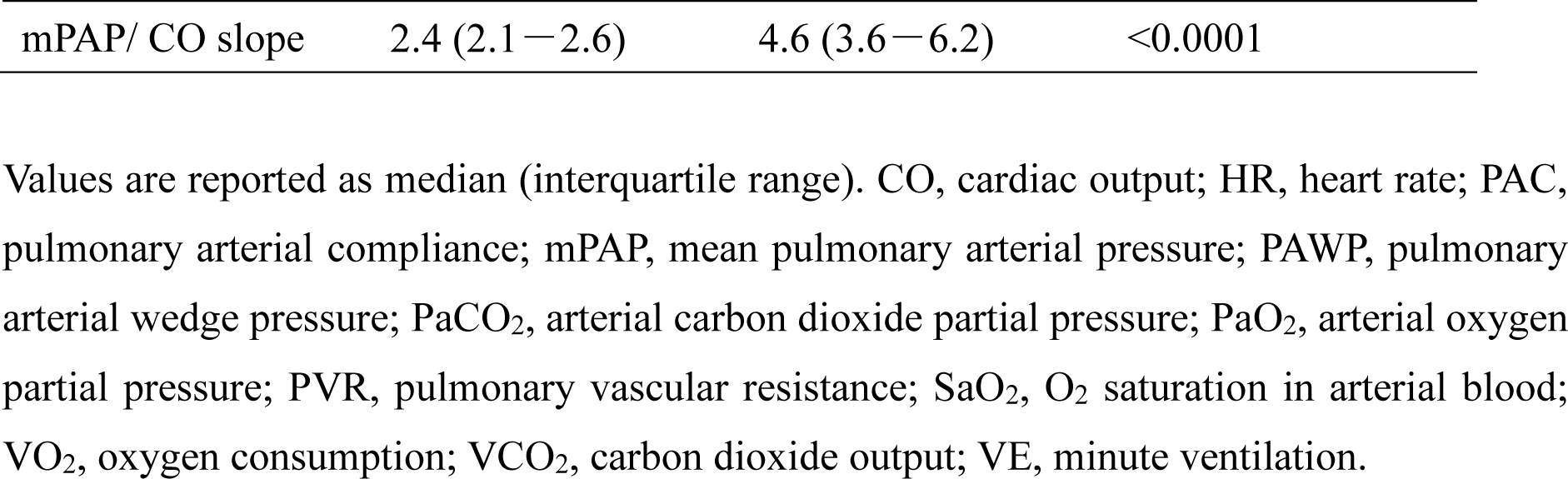
Comparison of Hemodynamic parameters, cardiopulmonary function, and oxygenation at CPET between Non-Ex-PH and Ex-PH.

## Notes

### Competing Interest Statement

The authors have declared no competing interest.

### Funding Statement

No conflicts of interest or financial disclosures are associated with this study.

### Author Declarations

This study complied with the Declaration of Helsinki and was approved by the Committee for Clinical Studies and Ethics of Kyorin University School of Medicine (Approval No.: H30-138).

